# Diastolic dysfunction in aortic stenosis is associated with greater damping of left ventricular recoil, and not myocardial stiffness

**DOI:** 10.1101/2024.09.20.24313309

**Authors:** Katrin Salman, Martin G Sundqvist, Jonathan Stevenson, Peter A Cain, Martin Ugander

## Abstract

**Background:** Aortic stenosis (AS) is associated with increasing severity of diastolic dysfunction as measured by conventional echocardiographic grading. The parameterized diastolic filling (PDF) method can describe diastolic function mechanistically, by analyzing filling using the physics of classical mechanics for spring recoil. The study aimed to use the PDF method to describe the mechanics of how diastolic dysfunction manifests in AS.

**Methods:** Patients (n=73) undergoing echocardiography were included according to AS of varying severity (normal/mild AS: maximum Doppler velocity across aortic valve (Vmax) <3.0 m/s, moderate/severe AS: Vmax≥3.0 m/s). PDF analysis of pulsed wave Doppler transmitral E-waves was performed using freely available software.

**Results:** Compared to normal/mild AS (n=41), patients with moderate/severe AS (n=32) had a left ventricle with a greater interventricular septal thickness (p=0.02) and higher E/e’ (p=0.007), but similar left ventricular ejection fraction (p=0.10) and left atrial volume index (p=0.21). PDF analysis showed that moderate/severe AS did not differ in myocardial stiffness (p=0.70), but had a higher myocardial damping (p=0.02), higher load (p=0.04), longer derived time constant of isovolumetric pressure decay (tau, p=0.004), higher filling energy (p=0.02), higher peak driving (p=0.02) and resistive (p=0.004) force of filling, lower kinematic filling efficiency index (p<0.001), but no difference in the load-independent index of diastolic function (p=0.62).

**Conclusions:** AS was primarily associated with a greater damping of LV recoil (increased viscoelasticity) and load, but without a change in myocardial stiffness. This provides novel insight into the mechanics of how diastolic dysfunction manifests in AS.

## Background

Aortic stenosis (AS) is the most common valvular lesion worldwide (1, 2), and it is known to contribute to the development of diastolic dysfunction (DD). Also, DD is an important risk factor for morbidity and mortality (1), and may both have a prognostic role and help identify high-risk patients (1, 2). The understanding of diastolic dysfunction is therefore of clinical importance. Currently, echocardiography is the most common clinical tool for assessing LV diastolic function. The 2016 guidelines (3) for evaluation of diastolic function provide a standardized scheme for diagnosis and grading of DD and offers a phenomenological classification based on categorizing echocardiographic measurements associated with impaired filling.

However, conventional echocardiographic grading of DD does not offer insight into the physical mechanistic manifestation of DD. Diastolic function assessment by echocardiography can also be analyzed using the parameterized diastolic filling (PDF) method. The PDF method entails describing early diastolic filling using the physics of classical mechanics for spring recoil, thereby offering a mechanistic description of diastolic LV filling. Software for analysis using the PDF method is freely for academic use in clinical research (4), and clinical normal reference values have been described (5). The PDF method has also been used to describe the mechanics of how diastolic dysfunction manifests in amyloidosis (6), hypertension (7), diabetes (8), and prognosis in heart failure (9).

The PDF method can uniquely provide insights into how the differing mechanistic manifestation of diastolic dysfunction varies between different disease states. Specifically, the PDF method can differentiate between changes in myocardial stiffness and damping. Stiffness contributes to the forces *driving* LV diastolic filling, and consequently is thought to be related to changes in titin, collagen, and recoil of the great vessels and pericardium. By comparison, damping contributes to the forces *opposing* LV diastolic filling, and consequently is thought to be related to changes in calcium sequestration and cross-bridge uncoupling. Thus, characterisation of the mechanics of diastolic dysfunction using the PDF method provides an opportunity to potentially identify pathophysiological mechanisms and ultimately targets for tailored pharmacological therapy that cannot otherwise be identified using conventional DD grading.

To our knowledge, no previous study has been performed using the PDF method to evaluate diastolic function in aortic stenosis. Therefore, the aim of this study was to improve the understanding of the physical mechanisms of diastolic dysfunction in AS by using the PDF method to describe the mechanics of how diastolic dysfunction manifests in AS.

## Methods

### Study population

This study was an observational cross-sectional prospective study with data collected from patients undergoing clinical transthoracic echocardiography at GenesisCare, Wesley Hospital, Brisbane, Australia, between August and December 2015. This study was approved by the UnitingCare Health Human Research Ethics Committee (Ref: 2024.02.396) with a retrospective waiver of individual subject informed consent. The patients were both in- and out-patients. Exclusion criteria were poor image quality, fusion of the E and A wave making the E wave not discernable for analysis, and severe stenosis or regurgitation in a valve other than the aortic valve. Patients were categorized according to AS of varying severity based on maximum Doppler velocity across the aortic valve <3.0 m/s for normal/mild AS, or ≥3.0 m/s for moderate/severe AS.

### Image acquisition and analysis

Baseline clinical and anthropometric data was noted in the medical records at the time of the echocardiography examination. Image acquisition was performed by experienced sonographers and was collected according to clinical guidelines for all conventional echocardiographic measurements (3). For the acquisition of E-waves for PDF analysis, the sample volume was positioned at the top at the mitral leaflet as per the normal acquisition for pulsed wave Doppler of transmitral flow to measure the E/A ratio. E-waves were collected during free breathing. Analysis of the E-waves for PDF parameters was performed using freely available software for PDF analysis (4). A comprehensive description of this methodology has previously been published (4). In summary, for each E-wave, a unique combination of the parameters for damping or viscoelasticity (c), stiffness (k), and load (x_0_) describe the shape of the E-wave (8, 10–12) thus describing different properties of left ventricular filling. The remaining parameters were calculated as previously described, namely damping index (c²-4k) (9), derived time constant of isovolumetric pressure decay (tau) (13) kinematic filling efficiency index (KFEI) (11), peak driving force of filling (kx_0_) (12), peak resistive force of filling (cV_max_) (12), filling energy (½kx ^2^) (8), and the load-independent index of diastolic filling (M) and intercept beta (12). All analysis was performed blinded to patient group.

### Statistical analysis

Statistical analysis was performed in SPSS (IBM SPSS Statistics for Windows, Version 24.0. Armonk, NY, USA). All data was tested for statistical normality using Kolmogorov-Smirnoff test. To assess differences between the groups the independent Mann-Whitney U test was used for the non-normally distributed data, and Student’s t-test for the normally distributed data. Data are presented as mean ± SD or median (interquartile range) as appropriate. Pearson’s chi-squared test was used to compare prevalence. A p-value <0.05 was considered statistically significant.

## Results

In total, 73 patients were included into either the normal/mild AS group (n=41, age 66.1±12.9 years, 41% female) or the moderate/severe AS group (n=32, age 75.1±11.0 years, 27% female). Baseline anthropometric and conventional diastolic measurement data are represented in Table 1. In summary, the group with moderate/severe AS expressed values consistent with a worse diastolic function measured with conventional parameters, while no other baseline data differed between the groups. The distribution of patients in the different diastolic dysfunction grading categories according to the 2016 American Society of Echocardiography guidelines are shown in Table 2. As expected, normal diastolic function was more prevalent among patients with normal/mild AS, while higher grade diastolic dysfunction was more common in moderate/severe AS. Figure 1 shows the results of E-wave analysis and the results from a representative subject from each group. The results of the PDF measurements are shown in Table 3. Patients with moderate/severe AS had a higher damping, load, derived time constant of isovolumetric pressure decay, potential energy of LV filling, and peak driving and peak resistive forces of filling. The kinematic filling efficiency index was lower for patients with moderate/severe AS, and the remaining PDF parameters did not differ between the groups.

**Table 1.** Baseline data and conventional diastolic measurements in patients with normal/mild and moderate/severe aortic stenosis.

|  | Normal/mild<br>(n=41) | Moderate/severe<br>(n=32) | p-value |
| --- | --- | --- | --- |
| Age, years | 66.1 ± 12.9 | 75.1 ± 11.0 | **0.002** |
| Weight, kg | 82.3 ± 19.0 | 81.1 ± 15.5 | 0.76 |
| Height, m | 173 (166-177) | 172 (160-177) | 0.21 |
| BMI, kg/m<sup>2</sup> | 27.8 ± 5.4 | 28.5 ± 4.8 | 0.58 |
| BSA, m<sup>2</sup> | 1.97 ± 0.26 | 1.94 ± 0.22 | 0.59 |
| Female sex, n (%) | 11 (27) | 13 (41) | 0.21 |
| Systolic blood pressure, mmHg | 132 ± 20 | 132 ± 30 | 0.94 |
| Diastolic blood pressure, mmHg | 80 (73-81) | 73 (66-80) | 0.09 |
| Heart rate, beats/minute | 65 (61-74) | 69 (62-77) | 0.27 |
| Left atrial volume index, ml/m<sup>2</sup> | 32.6 (27.1-39.9) | 37.6 (32.9-43.8) | 0.21 |
| Peak early diastolic transmitral flow velocity (E), m/s | 0.80 (0.60-0.90) | 0.70 (0.60-0.98) | 0.55 |
| Peak late diastolic transmitral flow velocity (A), m/s | 0.80 (0.60-0.90) | 0.90 (0.73-1.08) | **0.02** |
| Early to late diastolic transmitral flow velocity ratio (E/A) | 0.89 (0.79-1.23) | 0.80 (0.75-0.98) | 0.12 |
| Mean peak early diastolic myocardial velocity (e'), m/s | 0.085 (0.075-0.103) | 0.075 (0.051-0.084) | **0.002** |
| Peak early transmitral flow to peak early myocardial velocity ratio (E/e'), cm/s | 8.6 (7.1-11.2) | 10.0 (8.1-14.8) | **0.007** |
| Deceleration time (DT), ms | 217 (189-260) | 246 (210-310) | **0.03** |
| Ejection fraction, % | 63 (61-65) | 64 (61-69) | 0.10 |
| Interventricular septum thickness, mm | 10.0 (9.0-12.0) | 11.5 (10.0-13.0) | **0.02** |
| Left ventricular inner diameter, end diastole, mm | 46 (42-49) | 46 (44-50) | 0.78 |
| Estimated mean pressure gradient across aortic valve, mmHg | 8 (4-12) | 40 (29-57) | **<0.001** |
| Aortic valve maximum velocity, m/s | 2.0 (1.3-2.4) | 4.0 (3.5-4.9) | **<0.001** |

Data are presented as mean ± SD or median (interquartile range).

**Table 2.** Results of diastolic dysfunction grading according to the 2016 American Society for Echocardiography guidelines.

| Diastolic dysfunction grade | Total number, n=73 | Normal/mild, n=41 | Moderate/<br>severe, n=32 | p-value<br>Normal/mild vs<br>Moderate/severe |
| --- | --- | --- | --- | --- |
| Normal diastolic function, n (%) | 34 (47) | 25 (61) | 9 (28) | **0.005** |
| Indeterminate, n (%) | 23 (32) | 11 (27) | 12 (38) | 0.33 |
| Grade I, n (%) | 1 (1) | 1 (2) | 0 (0) | 0.34 |
| Grade II, n (%) | 13 (18) | 3 (7) | 10 (31) | **0.008** |
| Grade III, n (%) | 2 (3) | 1 (2) | 1 (3) | 0.86 |

**Figure 1.**
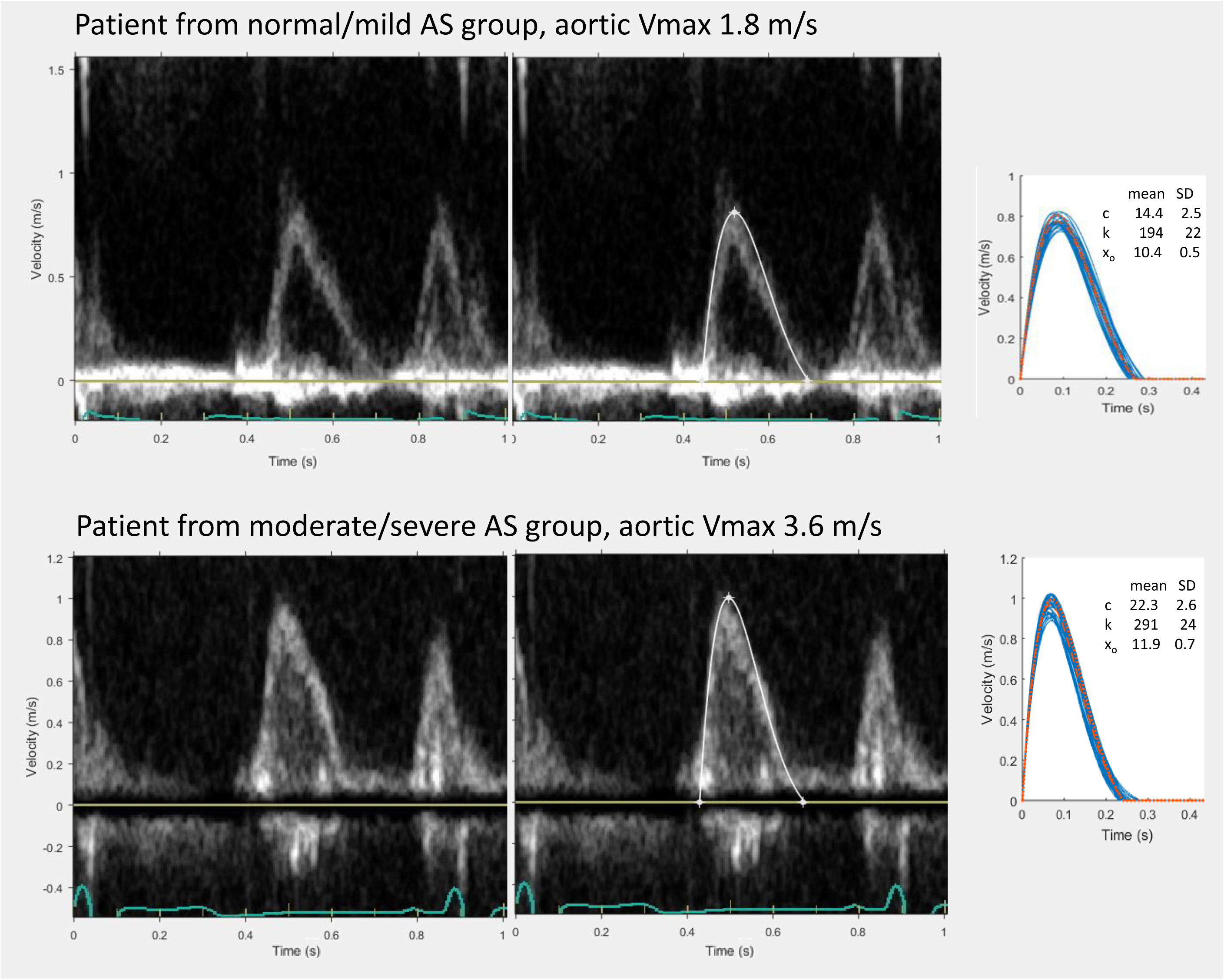
Semi-automatic curve fit of E-wave to determine PDF parameters. Top row: Patient from the normal/mild AS group with aortic Vmax 1.8m/s. Bottom row: Patient from the moderate/severe AS group with aortic Vmax 3.6m/s. Images show the transmitral Doppler velocities of E- and A-waves (left), semi-automatic curve fit of the E-wave after threshold detection (middle), and all analyzed E-waves for each patient collectively presented with mean values for c, k and x0.

**Table 3.**
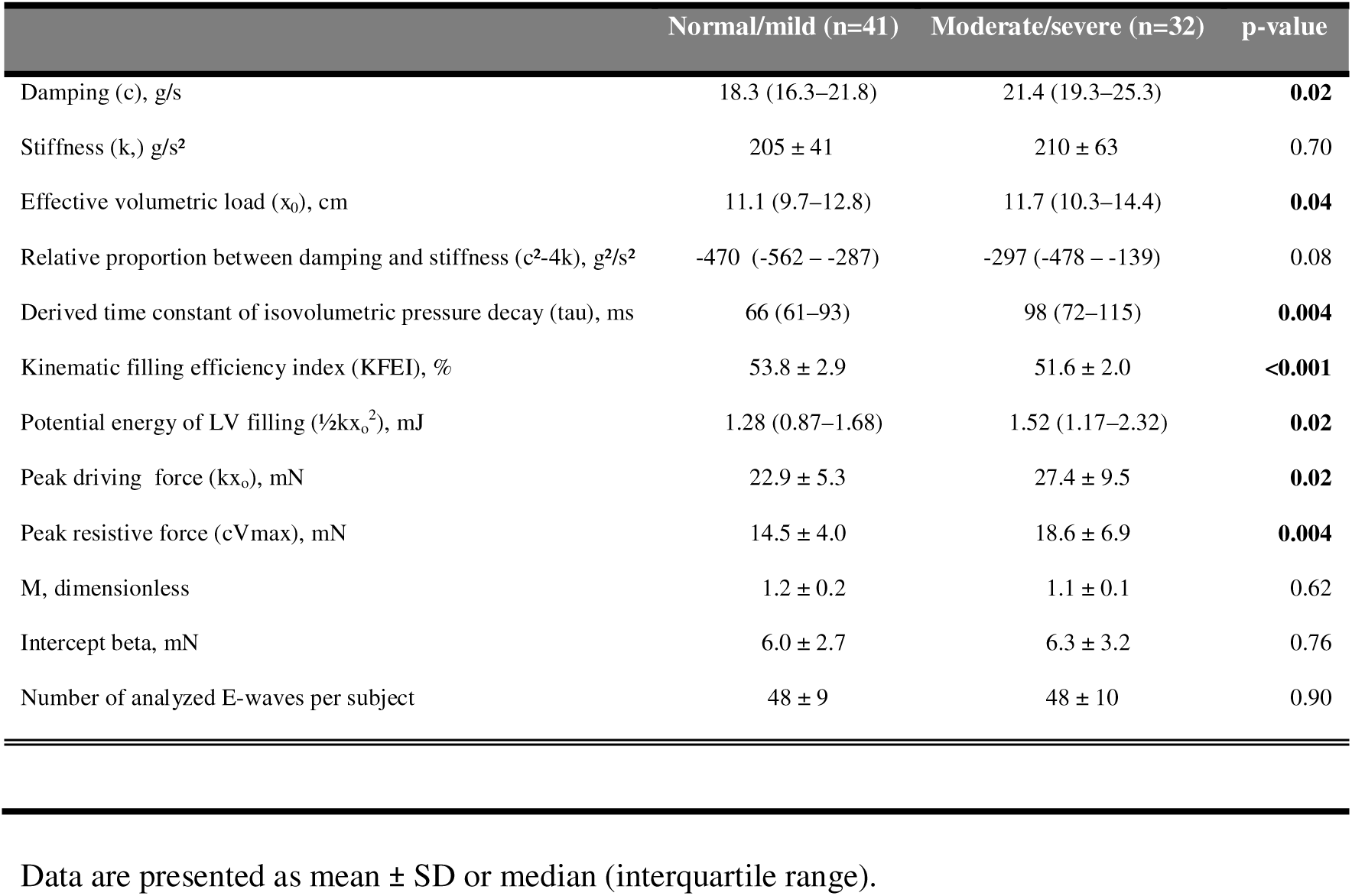
Results of PDF measurements for E wave analysis comparing normal/mild and moderate/severe aortic stenosis.

## Discussion

The current study is the first to investigate PDF measurements of diastolic function in patients with aortic stenosis. The major findings were that patients with a higher severity of aortic stenosis had a greater damping of left ventricular recoil and load, but no difference in myocardial stiffness.

### Conventional echocardiographic measures of diastolic function

Among the conventional echocardiographic parameters, both mitral flow measurements and tissue Doppler measurements differed between the groups in a way that was consistent with worse diastolic function in the patients with more severe AS. We found higher septal wall thickness in patients with moderate/severe AS, while there was no difference in EF. This is consistent with the concentric left ventricular remodeling that is known to be a compensatory consequence of chronic pressure overload in AS (1, 2). Nevertheless, even though we found a difference in septal wall thickness between our study groups, the magnitude of difference was subtle (1.5 mm). The conventional echocardiographic diastolic measurements in the moderate/severe AS group showed a higher late diastolic transmitral flow velocity (A-wave) and early transmitral flow to early myocardial velocity ratio (E/é), lower early diastolic myocardial velocity (é), and longer deceleration time, and these changes are all associated with worse diastolic function. Also, as expected, the patients in the moderate/severe AS group were older. There is a well-known association between age and diastolic dysfunction, with a decreasing E/A ratio, lower é and increasing E/é(14). Hence, one cannot rule out that these findings with regards to diastolic function may in part be related to age. Consistent with these findings, the moderate/severe AS group had a lower prevalence of normal diastolic function with regards to conventional diastolic dysfunction grading.

### PDF measures of diastolic function

For all investigated PDF parameters, the results for the normal/mild AS group largely fall within the previously published normal reference ranges (5).

#### Damping

Damping represents the energy loss or viscoelasticity of LV recoil, and thus reflects the resistive properties during LV filling. Damping was higher for the moderate/severe AS group. Similar findings have been reported for patients and rats with diabetes (8, 11, 15), as well as patients with hypertension (7). Among previous research, only one study has reported an increasing damping with age (16). In our study, age was higher in patients with more severe AS, but the blood pressure was equal between the groups. Age could therefore theoretically partly explain the measured difference in damping between the two groups. However, neither damping, stiffness nor load correlated with age among healthy individuals(5). Also, we did not have access to data regarding underlying diseases that could have affected the blood pressure such as smoking or treatment with antihypertensive drugs.

#### Stiffness

The PDF parameter stiffness is a measurement of LV rigidity, and we did not find differences in stiffness between the two groups of differing severity of aortic stenosis. While higher values of stiffness have been reported in women compared to men in a normal population (5), stiffness by PDF analysis has been found to not differ in patients with either hypertension (7) or diabetes (8). Stiffness can also be measured by invasive left heart catheterization in terms of dP/dV with the units mmHg/ml. Notably, the PDF measure of stiffness is closely and linearly correlated to invasively determined stiffness, and invasive stiffness can be calculated from the PDF measure of stiffness (17). In contrast to our results, previous invasive studies have found higher indices of stiffness in patients with aortic stenosis compared to controls (18, 19), and stiffness in aortic stenosis has been found to correlate with increasing wall thickness and/or myocardial cell diameter (18, 20). However, a straightforward comparison with regards to invasive stiffness cannot be made with our study since previous studies have used varying methods to define and measure stiffness. With that said, an explanation for our conflicting results regarding stiffness could be that the difference in severity of AS between the patients in our two study groups was not severe enough to give rise to a large enough thickening of the left ventricular chamber wall to impact stiffness.

Diastolic dysfunction in patients with AS is not a direct but rather an indirect consequence of remodelling of the heart. Although in most cases the degree of AS increases with time, the same severity of AS may have existed for different durations of time in different individuals.

#### Load

The effective volumetric load, x_0_, is the initial load-displacement at end-systole and is closely linearly related to the velocity time integral (VTI) of the E wave (VTI-E) (17). We found higher values for x_0_ in patients with moderate/severe AS indicating greater amount of blood entering the LV during early diastole. As far as we know, neither x_0_ nor VTI-E have been studied in patients with aortic stenosis. However, an increased ratio of the VTI between the E- and A-waves (VTI-E/A) has been studied and found to be associated with an increased risk of atrial fibrillation (21). In our study, the patients had no difference in LVEF, body surface area, and heart rate, and thus can be assumed to have similar cardiac output and stroke volume. Furthermore, subjects with similar stroke volumes would be assumed to have the same sum of VTI-E and VTI-A. An increased VTI-E in the presence of an unchanged stroke volume would result in a decreased VTI-A, and consequently an increased VTI-E/A. Thus, changes in load can be compared to previous studies of VTI-E/A in the literature. Several potential mechanisms that could lead to an increased VTI-E/A have been discussed, including reduced atrial function with preserved LV systolic function (18) . Taken together, the previous findings with regards to VTI-E/A agree broadly with our current findings with regards to increased load in the presence of decreased diastolic function and preserved LVEF. Also, atrial filling fraction, calculated as the ratio between VTI-A and the sum of VTI-E and VTI-A, has been reported to be lower in patients with severe AS, increased pulmonary wedge pressure, and reduced LVEF (22). In accordance with the reasoning above, a reduced atrial filling fraction with an unchanged stroke volume equates to a lower VTI-A and a higher VTI-E.

#### Potential energy, peak driving force, and peak resistive force

We found that the patients with more severe AS had a higher potential energy of LV filling and peak driving force of filling. This is expected, since these measurements are mathematically related to the products of load and stiffness, respectively, and load was found to be higher in our study. The peak driving force of filling is analogous to the peak atrioventricular pressure gradient, whereas the peak resistive force of filling is the opposing force counterbalancing the peak driving force, and is influenced by damping (23), and both were increased in our study. The potential energy of LV filling has been found to be higher in patients with hypertension (7), and the peak resistive force of filling has been found to be higher in patients with cardiac amyloidosis (6). This agreement is expected, since hypertension, amyloidosis, and AS all give rise to a thickened left ventricle wall that ultimately leads to diastolic dysfunction.

#### Damping-to-stiffness proportion

The proportion of contribution between damping and stiffness reflects the balance between these two parameters, and did not to differ between our two groups. Values of <-900 g²/s² have been found to predict one-year mortality in an elderly population with heart failure (9). Thus, we would have expected more negative values for patients with moderate/severe AS. On the contrary, similar to the findings in patients with cardiac amyloidosis (6), in both our study groups we found values that would fall within the normal reference ranges in a healthy population (5). Similarly to the finding regards to stiffness discussed above, this might indicate that the difference in the severity of AS between the groups was not large enough to impact this parameter.

#### Tau

In the group of moderate/severe AS, we found higher values of the time constant of isovolumetric pressure decay, tau. In our study, tau was calculated from PDF parameters as previously validated (17), and tau derived from PDF has been shown to be increased in impaired relaxation (13). Invasively determined tau, derived by analysis of the time constant of isovolumetric pressure decay for the left ventricle at catheterization, has been a frequently used metric to evaluate the relaxation properties of the left ventricular myocardium (24). Tau has been shown to be prolonged in patients with coronary artery disease, dilated cardiomyopathy, hypertrophic cardiomyopathy, and advanced age (24, 25). In a study of hemodynamics in AS, patients with severe AS were divided into subgroups based on LVEF and pulmonary wedge pressure, and found higher values of tau in patients with LVEF ≤55% and wedge pressure ≥15mmHg (22). They found an association between tau and both LV mass index and LV end-systolic volume index. Patients with severe AS and LV hypertrophy had diastolic relaxation abnormalities, but only those with depressed systolic function and a large increase in LV mass had prolonged LV isovolumetric pressure decay. They concluded that transmitral flow measures including the peak velocities of the E-wave, A-wave, the E/A ratio, and the atrial filling fraction are primarily determined by LA pressure, whereas isovolumetric relaxation depends on the severity of left ventricular hypertrophy and chamber dilatation (22). Those results agree with our results of higher values for tau and septal wall thickness in the higher severity AS group, even though our patients did not have reduced systolic function.

#### Load-independent index of diastolic filling

We did not find differences in the load-independent index of diastolic function, M, nor the intercept beta, when comparing patients with normal/mild and moderate/severe AS. These PDF measures are calculated after acquisition of E-waves with varying load induced by free breathing. M is the slope of the relationship between the change in peak driving force and peak resistive force. A higher peak resistive force for a given peak driving force will yield a lower M, and this has been shown to be associated with worse diastolic function (12). Also, the intercept beta is increased with worsened diastolic function (12). We would theoretically have expected M to be decreased and beta to be increased in the group with more severe AS, but this was not the case. This may be due to the fact that the severity of AS was not large enough between the groups in relation to the measurement variability of the measure.

#### KFEI

We found a lower kinematic filling efficiency index, KFEI, in patients with more severe AS. This index describes the efficiency of LV filling and is calculated as the ratio of the velocity time integral of the measured E-wave divided by the velocity time integral of a hypothetical E-wave with no resistance of filling, c=0, but the same measured stiffness and load. While KFEI has not previously been studied in AS, it has been shown to be reduced in patients with diabetes (11). Thus, our findings of a reduced KFEI are consistent with the expected impaired diastolic function associated with more severe AS.

#### The PDF method

The method of performing analysis according to the PDF method can vary, and influence measurement precision. We used freely availably software that previously has been used to analyze a median of 34 E-waves per patient, and showed good or excellent results with regards to interobserver variability for the majority of PDF parameters (coefficient of variance, range 2.5 – 18.7%) (4). In that study, measurement variability was higher than would be reasonably acceptable for two of the investigated PDF parameters, namely the load-independent index of diastolic filling, M, and intercept beta. Notably, in the current study we did not find any differences for either M or intercept beta between our study groups. This may be related to the limited measurement variability for these measures. In the current study, we analyzed on average 48 E-waves per patient, thus increasing the confidence in our measurements beyond results of previous studies that were limited to one or two E-waves per patient (4).

### Limitations

We did not have access to background information regarding the presence of clinical co-morbidities such as diabetes, hypertension, or medications that could have influenced the results, and this is a limitation. However, at the time of echocardiographic examination, the groups did not differ in systolic or diastolic blood pressure, suggesting no profound difference in hypertension. Furthermore, it would have been of interest to know the duration of aortic stenosis. Sample size is relatively modest but more than adequately powered to determine these mechanistic differences to gain physiological insights.

### Clinical implications

Our study indicates that diastolic dysfunction in patients with AS can be detected with PDF analysis. It is the first study to contribute to the mechanistic understanding of diastolic function in this patient group. Our results show that AS manifests with increased load and damping of filling, but not stiffness. Previous studies of hypertension, diabetes, and amyloidosis, have all shown higher values for damping compared to controls, but none have demonstrated an increased stiffness. Overall, our findings for patients with AS are broadly in agreement with previous findings regarding the mechanistic manifestation of diastolic dysfunction in both amyloidosis, diabetes and hypertensive heart disease, suggesting that these different pathophysiologies manifest similarly from a mechanistic perspective. A deeper understanding and better differentiation between stiffness and damping holds promise for identifiying more refined treatment options in diastolic dysfunction.

In conclusion, AS was primarily associated with a greater damping of LV recoil (increased viscoelasticity) and load, but without a change in myocardial stiffness. This provides novel insight into the mechanics of how diastolic dysfunction manifests in AS.

Conflicts of Interest: Nothing to Disclose.

## Data Availability

All data produced in the present study are available upon reasonable request to the authors.

